# Associations between governor political affiliation and COVID-19 cases, deaths, and testing in the United States

**DOI:** 10.1101/2020.10.08.20209619

**Authors:** Brian Neelon, Fedelis Mutiso, Noel T Mueller, John L Pearce, Sara E Benjamin-Neelon

## Abstract

**Introduction:** The response to the COVID-19 pandemic became increasingly politicized in the United States (US) and political affiliation of state leaders may contribute to policies affecting the spread of the disease. This study examined differences in COVID-19 infection, death, and testing by governor party affiliation across 50 US states and the District of Columbia.

**Methods:** A longitudinal analysis was conducted in December 2020 examining COVID-19 incidence, death, testing, and test positivity rates from March 15 through December 15, 2020. A Bayesian negative binomial model was fit to estimate daily risk ratios (RRs) and posterior intervals (PIs) comparing rates by gubernatorial party affiliation. The analyses adjusted for state population density, rurality, census region, age, race, ethnicity, poverty, number of physicians, obesity, cardiovascular disease, asthma, smoking, and presidential voting in 2020.

**Results:** From March to early June, Republican-led states had lower COVID-19 incidence rates compared to Democratic-led states. On June 3, the association reversed, and Republican-led states had higher incidence (RR=1.10, 95% PI=1.01, 1.18). This trend persisted through early December. For death rates, Republican-led states had lower rates early in the pandemic, but higher rates from July 4 (RR=1.18, 95% PI=1.02, 1.31) through mid-December. Republican-led states had higher test positivity rates starting on May 30 (RR=1.70, 95% PI=1.66, 1.73) and lower testing rates by September 30 (RR=0.95, 95% PI=0.90, 0.98).

**Conclusion:** Gubernatorial party affiliation may drive policy decisions that impact COVID-19 infections and deaths across the US. Future policy decisions should be guided by public health considerations rather than political ideology.

## Introduction

Coronavirus disease 2019 (COVID-19) has resulted in a global public health crisis. As of December 15, 2020, there have been over 16 million confirmed COVID-19 cases and 300,000 deaths in the US.^1^ In response to the pandemic, the governors of all 50 states declared states of emergency. Shortly thereafter, states began enacting policies to help stop the spread of the virus. However, these policies vary and are guided, in part, by decisions from state governors.

Through state constitutions and laws, governors have the authority to take action in public health emergencies. Earlier this year, nearly all state governors issued stay-at-home executive orders that advised or required residents to shelter in place.^2^ Recent studies found that Republican governors, however, were slower to adopt stay-at-home orders, if they did so at all.^3,4^ Moreover, another study found that Democratic governors had longer durations of stay-at-home orders.^5^ Further, researchers identified governor Democratic political party affiliation as the most important predictor of state mandates to wear face masks.^6^

Although recent studies have examined individual state policies, such as mandates to socially distance, wear masks, and close schools and parks,^3,4,6-8^ multiple policies may act together to impact the spread of COVID-19. Additionally, the pandemic response has become increasingly politicized.^7,9,10^ As such, political affiliation of state leaders, and specifically governors, might best capture the omnibus impact of state policies. Therefore, the purpose of this study was to quantify differences in incidence, death, testing, and test positivity over time, stratified by governors’ political affiliation among the 50 states and DC.

## Methods

A longitudinal analysis examined COVID-19 incident cases, death rates, polymerase chain reaction (PCR) testing, and test positivity from March 15 (March 24 for testing and test positivity) through December 15, 2020 for 50 states and DC. Based on prior studies,^3,4,6,7^ it was hypothesized that states with Democratic governors would have higher incidence, death, and test positivity rates early in the pandemic due to points of entry for the virus,^11,12^ but that the trends would reverse in later months, reflecting policy differences that break along party lines. The Institutional Review Boards at the Medical University of South Carolina and Johns Hopkins Bloomberg School of Public Health deemed this research exempt.

Governor party affiliation was documented for each US state; for DC, mayoral affiliation was used. Daily incident cases and deaths were obtained from the COVID Tracking Project.^13^ PCR testing and test positivity data came from the US Department of Health and Human Services.^14^ Potential confounders included state population density,^15^ census region,^15^ state percentage of residents aged 65 and older,^15^ percentage of Black residents,^15^ percentage of Hispanic residents,^15^ percentage below the federal poverty line,^15^ percentage living in rural areas,^16^ percentage with obesity,^17^ percentage with cardiovascular disease,^18^ percentage with asthma, percentage smoking,^9^ number of physicians per 100,000 residents,^16^ and percentage voting Democratic (versus Republican) in the 2020 presidential election.^19^

### Statistical analysis

Bayesian negative binomial models were used to examine incident case, death, testing, and test positivity rates. The models included penalized cubic Bsplines for the fixed and random temporal effects. Models adjusted for the above covariates. Ridging priors were assigned to the fixed and random spline coefficients.^20^ Posterior computation was implemented using Gibbs sampling.^18,21^ Model details, including prior specification, computational diagnostics, and sensitivity analyses appear in the online Appendix.

Models were stratified by governors’ affiliation, and posterior mean daily rates were graphed with their 95% posterior intervals (PIs). Adjusted risk ratios (RRs) and 95% PIs were calculated to compare states, with RRs > 1.00 indicating higher rates among Republican-led states. Analyses were conducted using R version 3.6 (R Core Team, 2019).

## Results

The sample comprised 26 Republican-led and 25 Democratic-led states. Figures 1(a-b) present incidence trends (cases per 100,000) and adjusted RRs by gubernatorial affiliation. Republican-led states had fewer cases from March to early June 2020. However, on June 3 the association reversed (RR=1.10, 95% PI=1.01, 1.18), indicating that Republican-led states had on average 1.10 times more cases per 100,000 than Democratic-led states. The RRs increased steadily thereafter, achieving a maximum of 1.77 (95% PI=1.62, 1.90) on June 28 and remaining positive for the remainder of the study, although the PIs overlapped 1.00 starting on December 3.

**Figure 1.**
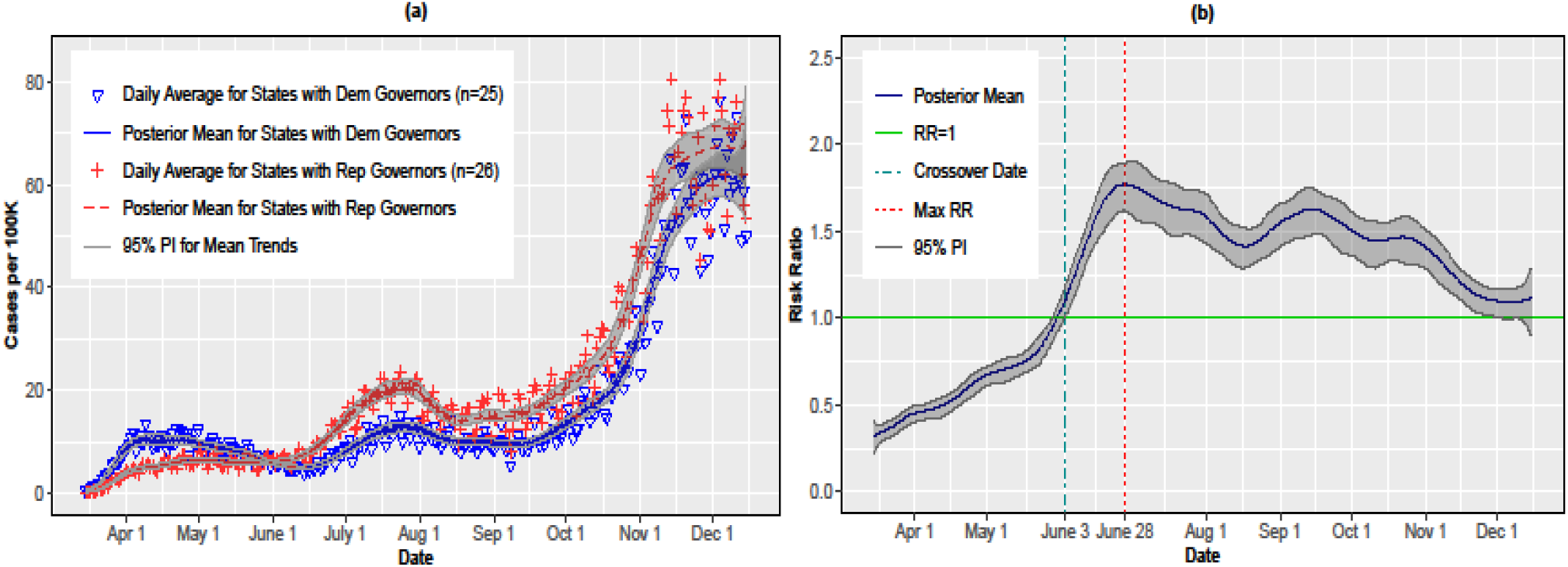
(a) Per capita COVID-19 incidence rates by governor affiliation; (b) adjusted risk ratios (RRs) and 95% posterior intervals (PIs). RRs > 1 indicate higher rates for Republican governors.

A similar pattern emerged for deaths shown in Figures 2(a-b). Republican-led states had lower death rates early in the pandemic, but the trend reversed on July 4 (RR=1.18, 95% PI=1.02,1.31). The RRs increased through August 5 (RR=1.80, 95% PI=1.57, 1.98) and the PIs remained above 1.00 until December 13 (RR=1.20, 95% PI=0.96, 1.39).

**Figure 2.**
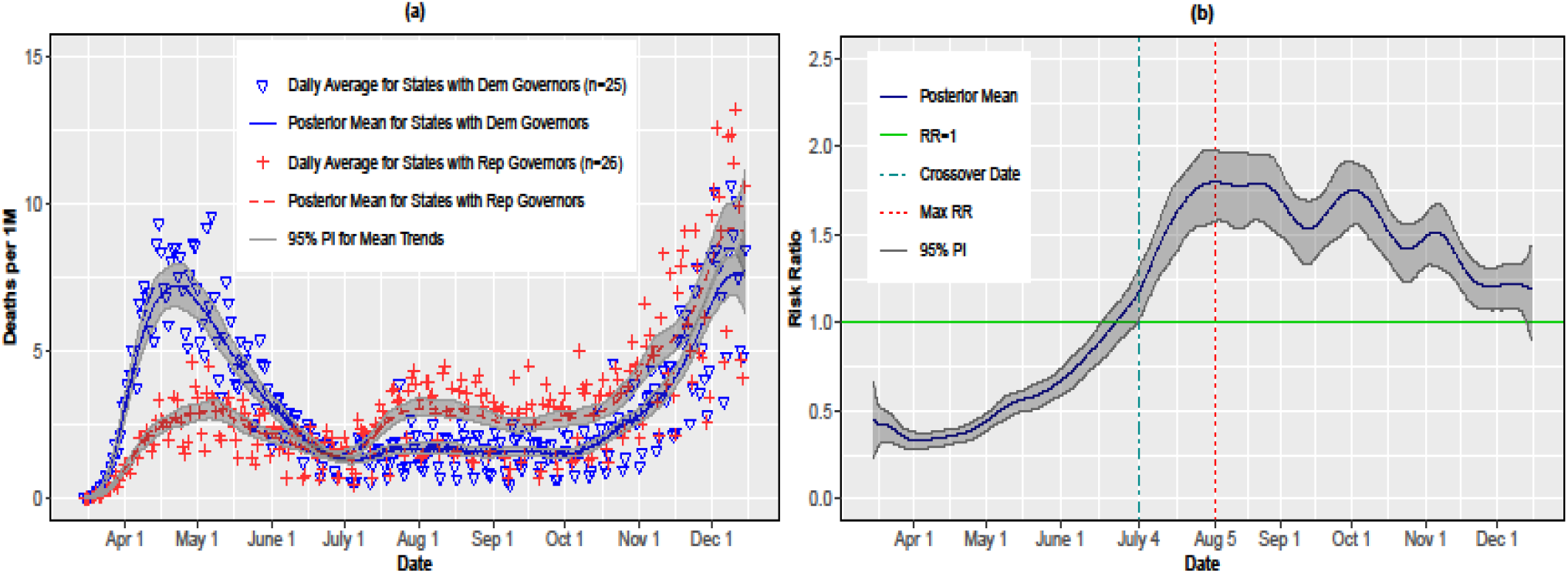
(a) Per capita COVID-19 death rates by governor affiliation; (b) adjusted risk ratios (RRs) and posterior intervals (PIs). RRs > 1 indicate higher rates for Republican governors.

Testing rates (Figures 3a-b) tracked similarly for Republican and Democratic states until September 30 (RR=0.95, 95% PI=0.90, 0.98). By November 27, the testing rate for Republican-led states was substantially lower than Democratic states (RR=0.77, 95% PI=0.72, 0.80). The test positivity rate (Figures 4a-b) was higher for Republican-led states starting on May 30, and was 1.70 (95% PI=1.65, 1.74) times higher on June 23.

**Figure 3.**
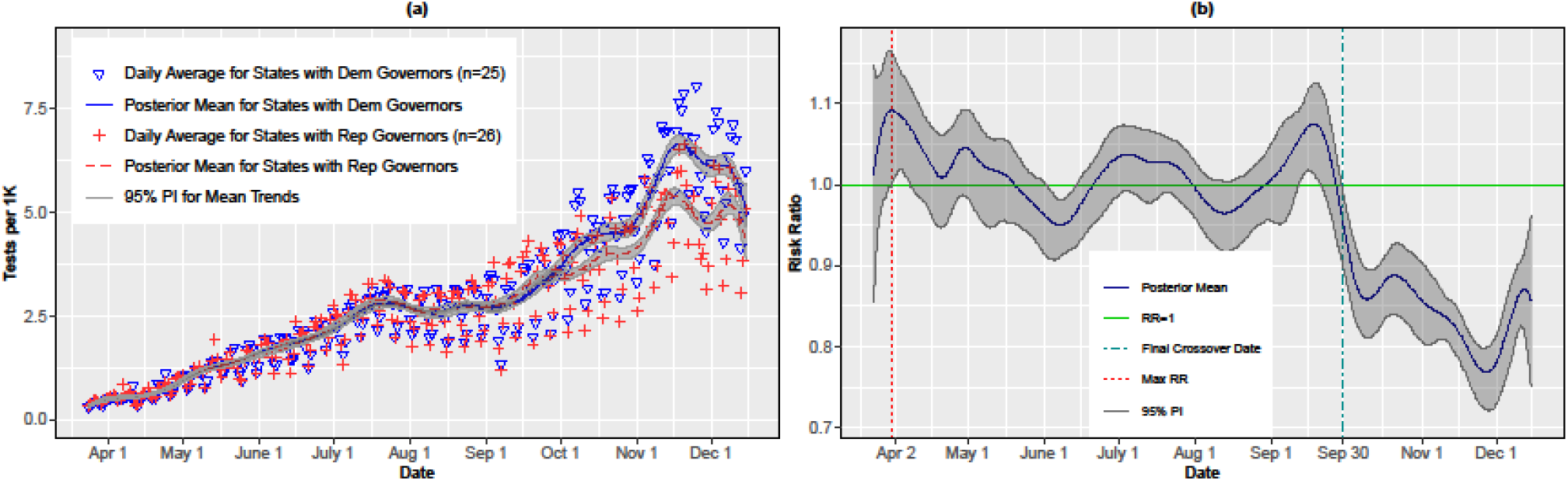
(a) Per capita COVID-19 testing rates by governor affiliation; (b) adjusted risk ratios (RRs) and posterior intervals (PIs). RRs > 1 indicate higher rates for Republican governors.

**Figure 4.**
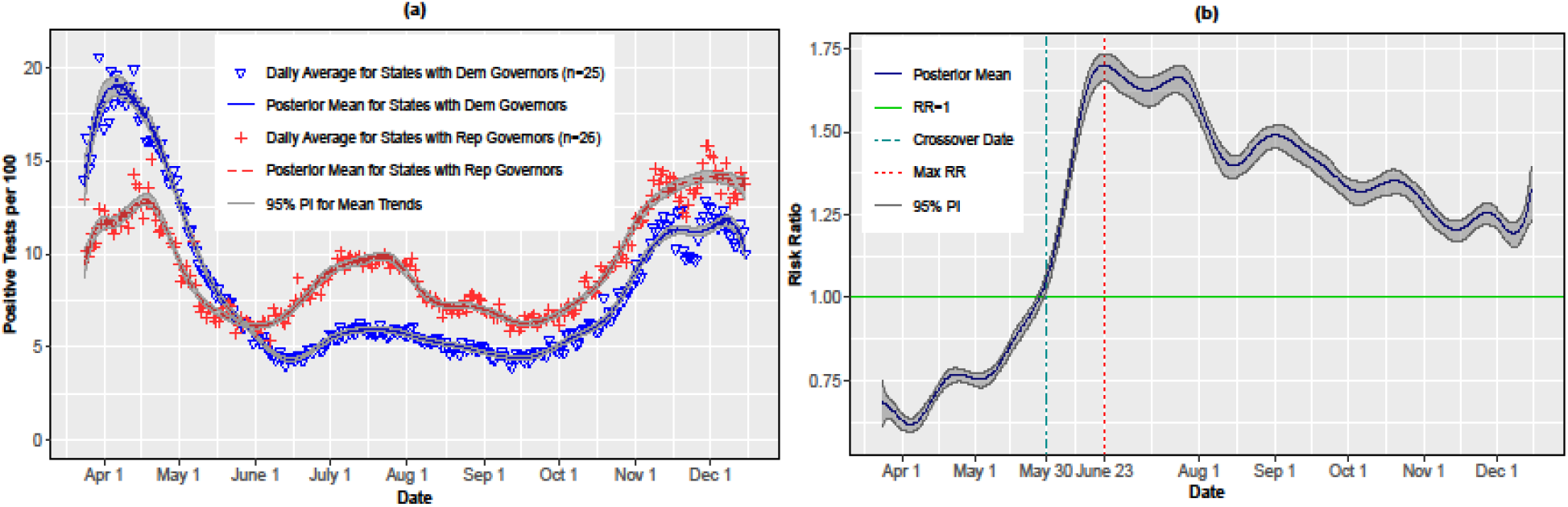
(a) COVID-19 test positivity rates per 100 tests by governor affiliation; (b) adjusted risk ratios (RRs) and posterior intervals (PIs). RRs > 1 indicate higher rates for Republican governors.

## Discussion

In this longitudinal analysis, Republican-led states had fewer per capita COVID-19 cases, deaths, and positive tests early in the pandemic, but these trends reversed in early May (positive tests), June (cases), and July (deaths). Testing rates were similar until September, when Republican states fell behind Democratic states. The early trends could be explained by high COVID-19 cases and deaths among Democratic-led states that are home to initial ports of entry for the virus in early 2020.^11,12^ However, the subsequent reversal in trends, particularly with respect to testing, may reflect policy differences that could have facilitated the spread of the virus.^3,4,6-9^

Adolph et al. found that Republican governors were slower to adopt both stay-at-home orders and mandates to wear face masks.^3,6^ Other studies have shown that Democratic governors were more likely to issue stay-at-home orders with longer durations.^4,5^ Moreover, decisions by Republican governors in spring 2020 to retract policies, such as the lifting of stay-at-home orders on April 28 in Georgia,^22^ may have contributed to increased cases and deaths. Democratic states also had lower test positivity rates from May 30 through December 15, suggesting more rigorous containment strategies in response to the pandemic. Thus, governors’ political affiliation might function as an upstream progenitor of multifaceted policies that, in unison, impact the spread of the virus. Although there were exceptions in states such as Maryland and Massachusetts, Republican governors were generally less likely to enact policies aligned with public health social distancing recommendations.^3^

This is the first study to quantify differences over time based on governor party affiliation. There are, however, limitations. This was a population-level rather than individual-level analysis. Although analyses were adjusted for potential confounders (e.g., rurality), the findings could reflect the virus’s spread from urban to rural areas.^11,12^ Additionally, as with any observational study, causality cannot be inferred. Finally, governors are not the only authoritative actor in a state; governors in states like Wisconsin may have been limited by Republican-controlled legislatures. Future research could explore associations between party affiliation of state or local legislatures, particularly when these differ from governors.

These findings suggest that governor political party affiliation may differentially impact COVID-19 incidence and death rates. Attitudes toward the pandemic were highly polarized in 2020.^7,9,10,23-25^ Future state policy actions should be guided by public health considerations rather than political expedience^26^ and should be supported by a coordinated federal response within the new presidential administration.

## Data Availability

All data used in this paper are publicly available.

## Acknowledgments

Dr. Neelon is a part-time employee of the Department of Veterans Affairs. The content of this article does not represent the views of the Department of Veterans Affairs or the U.S. government. The article represents the views of the authors and not those of the VA or Health Services Research and Development. Dr. Mueller was supported by the National Heart, Lung, And Blood Institute of the National Institutes of Health under Award Number K01HL141589 (PI: Mueller). The funder had no influence on the study design, implementation, or findings. Dr. Neelon had full access to all data in the study and takes responsibility for the integrity of the data and the accuracy of the data analysis. Dr. Neelon, Dr. Mueller, Dr. Pearce and Dr. Benjamin-Neelon contributed to the concept and design of the study. Dr. Neelon and Mr. Mutiso contributed to acquisition, analysis, and interpretation of the data. Dr. Neelon and Benjamin-Neelon drafted the manuscript, and all Dr. Mueller, Dr. Pearce, and Mr. Mutiso provided critical revisions. A preprint of this manuscript is posted on *MedRχiv* at https://www.medrxiv.org/content/10.1101/2020.10.08.20209619v1. No financial disclosures were reported by the authors of this manuscript.

